# The combination of aggressive hydration and rectal non-steroidal anti-inflammatory drugs is not superior to rectal non-steroidal anti-inflammatory drugs alone in the prevention of pancreatitis after endoscopic retrograde cholangiopancreatography: a systematic review and meta-analysis

**DOI:** 10.1101/2022.12.02.22283014

**Authors:** Xiang Cheng, Feixiang Yang, Xingxin Yang, Ning Zhang, Xiaoming Li, Bo Chen

**Affiliations:** Department of Gastroenterology, The First Affiliated Hospital of Anhui Medical University, Hefei 230022, China; First School of Clinical Medicine, Anhui Medical University, Hefei 230032, China; Department of Medical Psychology, School of Mental Health and Psychological Sciences, Anhui Medical University, Hefei 230032, China

**Keywords:** Anti-Inflammatory Agents, Non-Steroidal, Cholangiopancreatography, Endoscopic Retrograde, Fluid Therapy, Meta-Analysis, Systematic Review

## Abstract

**Background:** endoscopic retrograde cholangiopancreatography (ERCP) can lead to many high-risk complications, of which acute pancreatitis is the most prevalent and serious one. Whether patients who receive prophylactic rectal non-steroidal anti-inflammatory drugs (NSAIDs) need to be combined with aggressive hydration remains controversial.

**Aim:** The study was performed to determine whether there is collaborative facilitation between rectal NSAIDs and aggressive hydration in preventing post-ERCP pancreatitis (PEP).

**Methods:** We searched all eligible studies on the preventive effects of active rehydration and NSAIDs on PEP from multiple databases including ClinicalTrials, PubMed, CQVIP, Embase, Web of Science, CNKI, Cochrane Library, and Wanfang Data. We performed a meta-analysis of the data related to the incidence of PEP as well as the serious cases including the incidence of severe PEP and mortality.

**Results:** This meta-analysis included three published studies of randomized controlled trials with 1110 patients. Our results showed that additional aggressive hydration was not significantly effective for the prevention of PEP in patients who were already receiving rectal NSAIDs (odds ratio [OR], 0.43; 95% confidence interval [CI], 0.12-1.57; P=0.20). With regard to the prevention of serious cases, compared with rectal NSAIDs alone, aggressive fluid hydration combined with rectal NSAIDs did not reduce the morbidity of severe PEP (OR, 0.32; 95% CI, 0.10-1.08; P=0.07), nor did it reduce overall mortality (OR, 0.81; 95% CI, 0.28-2.36; P=0.70).

**Conclusion:** Aggressive perioperative hydration combined with rectal NSAIDs was not superior to rectal NSAIDs along in the prevention of PEP.

## Introduction

Endoscopic retrograde cholangiopancreatography (ERCP) is conducted to evaluate the pathological changes in bile duct, pancreatic duct, and hepatopancreatic ampullae, such as stones, tumors, and inflammation [1]. The improvement of endoscopic technology advances the application of ERCP in the diagnosis and management of pancreatic and biliary tract diseases, making ERCP a major diagnostic and therapeutic intervention for biliopancreatic lesions [2]. Post-ERCP pancreatitis (PEP) is the most prevalent and serious ERCP-associated complication, with an incidence that ranges between 3% and 5% in low-risk patients and as high as 15% in high-risk patients [1-4]. ERCP induces pancreatic injury and inflammation in two main mechanisms: mechanical damage from instrument handling and hydrostatic compression damage from contrast media [5, 6]. There are many ERCP procedures involving prolonged instrument operation of the pancreatic duct, such as long-term or repeated pancreatic intubation, which can cause direct injury to the pancreatic duct or ampulla. In addition, some procedures operated by an electric knife can result in mechanical and thermal injury, subsequently lead to secondary trypsin injury after reactive edema and obstruction of the pancreatic duct [5]. The pancreatic injury induced by contrast mainly includes hydrostatic compression damaged by overuse of contrast or chemical or allergic damage induced by contrast [5-7]. Although George *et al*. found that no remarkable difference existed in the incidence of PEP between high and low-osmolality contrast, the role of contrast in pancreatic damage remains controversial [8]. Other underlying mechanisms include zymogen activation, genetic defects, and flora disorder, however, these do not affect as large as mechanical damage or contrast damage to pancreatic injury [9].

PEP is defined as the elevation of serum amylase level more than three times the upper limit of normal within 24 hours, accompanied by typical clinical symptoms of pancreatitis, such as persistent pancreatitis-like abdominal pain [10]. There are many academic discussions on effective strategies to prevent PEP, among them, pancreatic duct stenting and perioperative rectal nonsteroidal anti-inflammatory drugs (NSAIDs) are widely accepted [11-13]. Additionally, aggressive hydration has been also recommended by clinical researchers due to its promising preventive effect [14, 15]. The mechanism of NSAIDs in preventing acute pancreatitis is related to their potent inhibitory effects on cyclooxygenase, phospholipase A2, and interaction between neutrophils and endothelial cells, which activate inflammatory cascade and lead to pancreatitis [16]. NSAIDs are an attractive option to prevent PEP because of their affordability and easy patient management [17]. There is evidence that aggressive perioperative hydration presents satisfactory efficacy in prevention of PEP [18-23]. Hydration improves blood perfusion and stabilizes the pancreatic microcirculation by attenuating tissue acidification, improving the prognosis of patients [24-27]. The mechanism of hydration is to protect the pancreatic microcirculation and NSAIDs are aimed at suppressing the inflammatory response, however, whether hydration and rectal NSAIDs exhibit synergistic effects remains unclear [28-31]. The discussion on this issue is still controversial [17, 32]. Therefore, we comprehensively reassess the efficacy of aggressive hydration with rectal NSAIDs in comparison with rectal NSAIDs alone, providing evidence-based support for the prevention of PEP.

## Methods

### Search strategy

The study comprehensively searched ClinicalTrials, PubMed, CQVIP, Embase, Web of Science, CNKI, Cochrane Library, and Wanfang Data, and searched for all relevant articles published before September 2021. After several attempts, we chose to sacrifice a little precision to ensure the comprehensiveness of the search, while comparing several meta-analyses related to the content of this study, and finally determined the search formula for this study. The search formula applied in PubMed is as follows: (post-ercp [All Fields] AND (“pancreatitis” [MeSH Terms] OR “pancreatitis” [All Fields]) AND (“prevention and control” [Subheading] OR (“prevention” [All Fields] AND”control” [All Fields]) OR”prevention and control” [All Fields] OR “prevention” [All Fields])) AND (randomized controlled trial [Publication Type] OR randomized [Title/Abstract] OR placebo [Title/Abstract]).

### Inclusion and exclusion criteria

We included all randomized controlled trials that contained a comparison of the efficacy of rectal NSAIDs and rectal NSAIDs in combination with active fluid hydration for PEP prevention. Our inclusion criteria included the following: a) RCTs that directly compared rectal NSAIDs combined with aggressive hydration and rectal NSAIDs alone; b) our primary outcome was the rate of PEP and secondary outcomes were the rate of severe and fatal cases of PEP (PEP was defined as serum amylase levels three times higher than the upper limit of normal within 24 hours after ERCP and persistent pancreatitis-like abdominal pain was observed). We excluded the following: a) RCTs in relation to combination of rectal NSAIDs and pancreatic duct stents or RCTs that did not contain aggressive hydration; b) studies that did not report complete experimental data or whose full text were not available; c) not RCTs.

### Data extraction and management

We determined a data extraction table after several tests in advance. Two independent reviewers extracted data from the included literature using the unified form. The main information extracted from the data extraction form included: lead author, year of publication, patient demographics (ie, country, mean age, sex), sample size, intervention measure (ie, type of NSAIDs, NSAIDs dose, type of infusion fluid, duration and volume of infusion), prevalence of PEP, mortality, and incidence of adverse effects for each study. We also extracted relevant patient metrics and complication data that may be useful for each experiment. In case of disagreement between two reviewers, the third reviewer negotiated and resolved it. All reviewers agreed at the final results.

### Risk of bias assessment

We evaluated the risk of bias in the relevant literature according to the seven aspects included in the Cochrane Handbook for Systematic Reviews of Interventions, including publication bias, time lag bias, multiple publication bias, publication position bias, citation bias, language bias, and outcome reporting bias. We also assessed the quality of the evidence based on GRADE ratings, which evaluate evidence quality depending on study limitations, inconsistency of results, indirectness of evidence, imprecision of results, and reporting bias. We classified the quality of evidence into four levels (high, moderate, low, or very low).

### Statistical analysis

Statistical analysis was performed by RevMan software (version 5.4). Data were collected with dichotomous variables, and odds ratios (OR) and 95% confidence intervals (CI) were calculated. Statistical heterogeneity between studies was measured by I^2^, and significant heterogeneity was indicated when I^2^ > 50%. If the heterogeneity is not significant, a model or a random-effects model is generally employed. Conversely, only the fixed-effects model is used while the heterogeneity is significant. We performed statistical analysis of studies data using the random-effects model in this meta-analysis. Statistical significance was considered when the p-value < 0.05. Although the number of included literatures was small, a sensitivity analysis was still performed. Sensitivity analysis showed that our results were not affected by the heterogeneity of different literatures.

## Results

### Search results

1355 relevant articles were found from ClinicalTrials, PubMed, CQVIP, Embase, Web of Science, CNKI, Cochrane Library, and Wanfang Data. We showed the complete retrieval process and screening details in **Figure 1**. After multiple screening, literatures not included in the inclusion criteria or with missing data were excluded. Finally, three RCTs with 1110 patients were included in this study and related results were presented in **Figures 1-5** and **Table 2**. The PRISMA statement was followed in the review process of our systematic review [33].

**Figure 1.**
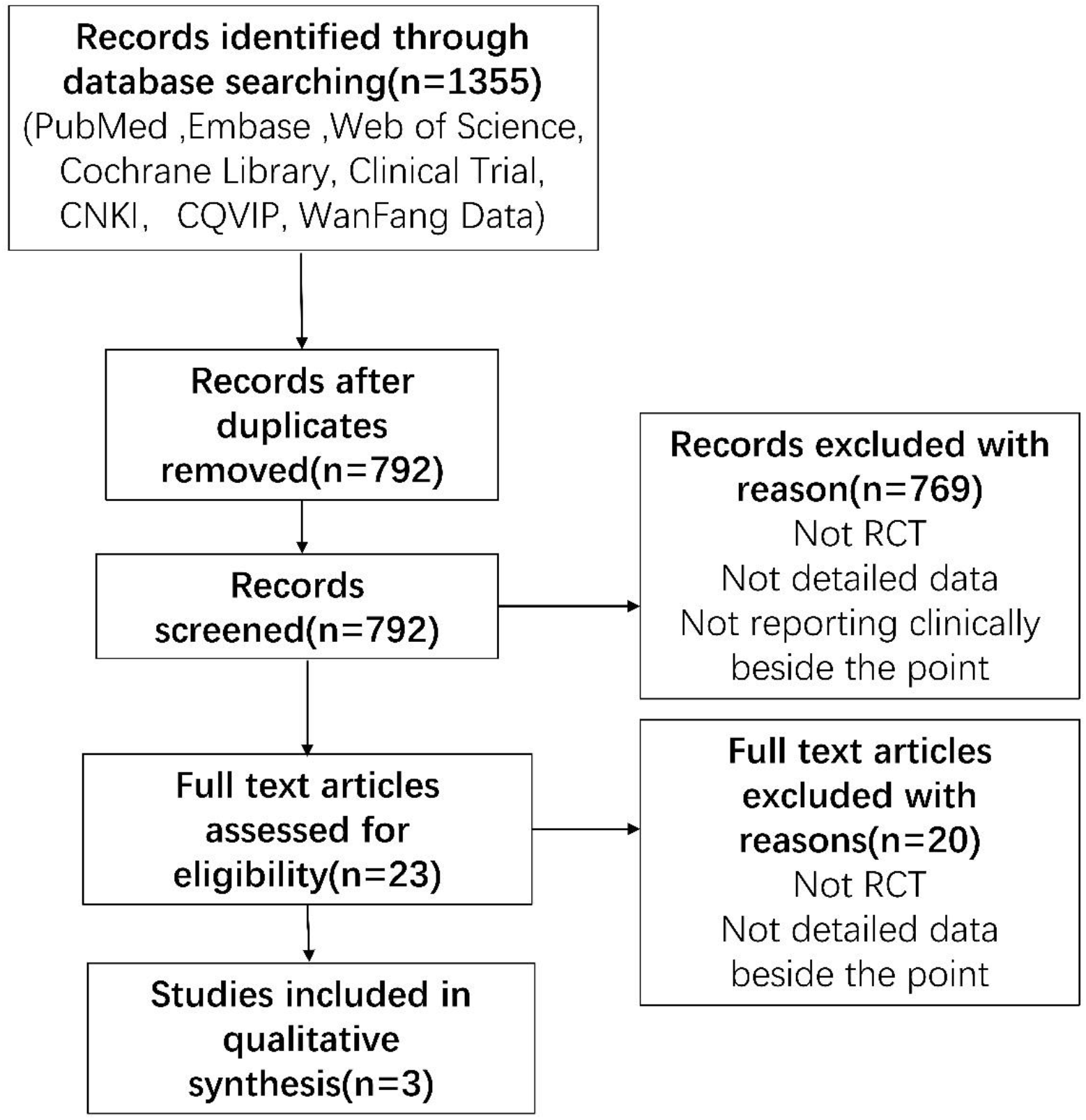
The PRISMA flow diagram of selected studies.

### Characteristics of trials

We included 3 published studies of RCTs with 1110 patients. Of these three RCTs, two studies were in the United States [34, 35] and one study was in Iran [17]. One was published in 2021, one in 2017, and one in 2016, all relatively novel studies within five years of each other and of good reference value. The three included studies were full-text papers. With respect to hydration reagent, in one study, the experimental group was injected with normal saline, while the control group did not [17]; and in the other two studies [34, 35], Lactate Ringer solution was used in experimental group, while normal saline was used in control group. For NSAIDs, two trials were administered indomethacin [17, 34], and one trial was administered indomethacin or diclofenac randomly [35]. All trials were administered rectally at a dose of 100mg. More details such as patient information, interventions were presented in **Table 1**.

**Table 1.** Characteristics of included studies

| Study name | Weiland *et al.* | Mok *et al.* | Hosseini *et al.* |
| --- | --- | --- | --- |
| Location | USA | USA | Iran |
| Year | 2021 | 2017 | 2016 |
| Patient Age, years | ≤40 years old, 81 (AH)/67(SH)<br>>40 years old, 307(AH)/358(SH) | Mean Age 63(AH)/62(SH) | Mean ± SD<br>47.91 ± 11.06(AH)/<br>51.20 ± 12.12 (SH) |
| Sex | Female,232(AH)/250(SH)<br>Male,156(AH)/175(SH) | Female,23(AH)/33(SH)<br>Male,25(AH)/15(SH) | Female,62(AH)/40(SH)<br>Male,39(AH)/60(SH) |
| Previous pancreatitis | 13(AH)/13(SH) | 0(AH)/1(SH) | - |
| Pancreatic duct stent placement | 58(AH)/73(SH) | 10(AH)/12(SH) | - |
| Difficult cannulation | 113(AH)/123(SH) | 6(AH)/8(SH) | - |
| Trainee involvement | 33(AH)/34(SH) | 48(AH)/48(SH) | - |
| Study design | RCT n=813(388AH/425SH) | RCT n=96(48AH/48SH) | RCT<br>n=201(101AH/100SH) |
| Intervention (AH/SH) | **AH:** Receive rectal NSAIDs 100mg within 30 min before or after ERCP procedure and 20 mL/kg of Lactate Ringer solution intravenously within 60 minutes as the beginning of the procedure, followed by 3 mL/kg every hour for 8 hours.<br>**SH:** Receive rectal NSAID 100 mg alone and restrict intravenous infusion of normal saline. | **AH:** Received preoperative rectal injection of indomethacin 100 mg and preoperative intravenous 1000 ml of Lactate Ringer solution.<br>**SH:** Received intraoperative intravenous normal saline and preoperative rectal injection of indomethacin 100 mg. | **AH:** rectal indomethacin and intravenous normal saline.<br>**SH:** 100 mg of Indomethacin rectally two hours before the ERCP procedure. |

**Table 2.** Assessment of quality of evidence.

| Outcomes | Illustrative comparative risks*(95%CI) |  | Relative effect (95% CI) | No of Participants(studies) | Quality of the evidence (GRADE) |
| --- | --- | --- | --- | --- | --- |
|  | Assumed risk | Corresponding risk |  |  |  |
|  | NSAIDs alone | Fluid hydration plus NSAIDs |  |  |  |
| PEP | Study population |  | OR 0.43<br>(0.12 to 1.57) | 1110(3 studies) | ⊕ ⊕ ⊕ ⊖<br>moderate |
|  | 98 per 100 | 45 per 1000<br>(13 to 145) |  |  |  |
|  | Moderate |  |  |  |  |
|  | 110 per 1000 | 50 per 1000<br>(15 to 163) |  |  |  |
| Severe PEP | Study population |  | OR 0.32<br>(0.10 to 1.08) | 909 (2studies) | ⊕ ⊖ ⊖ ⊖<br>very low |
|  | 23 per 1000 | 8 per 1000<br>(2 to 25) |  |  |  |
|  | Moderate |  |  |  |  |
|  | 22 per 1000 | 7 per 1000<br>(2 to 24) |  |  |  |
| Death | Study population |  | OR 0.81<br>(0.28 to 2.36) | 909 (2studies) | ⊕ ⊖ ⊖ ⊖<br>very low |
|  | 17 per 1000 | 14 per 1000<br>(5 to 39) |  |  |  |
|  | Moderate |  |  |  |  |
|  | 28 per 1000 | 23 per 1000<br>(8 to 64) |  |  |  |

CI = Confidence interval, OR = Odds ratio, PEP = post -ERCP pancreatitis.

### Risk of bias in included studies

Among three RCTs, one study [34] was single-blinded study. Two studies [17, 35] were double blinded studies. One study [35] did not provide enough information about incomplete outcome data (attrition bias), while another two studies [17, 34] provided complete data in incomplete outcome data. Funnel plots cannot assess true asymmetry when the sample size is less than 5 studies, so funnel plots were not used to assess publication bias [36]. The assessment of publication bias of the included studies was shown in **Figure 2**.

**Figure 2.**
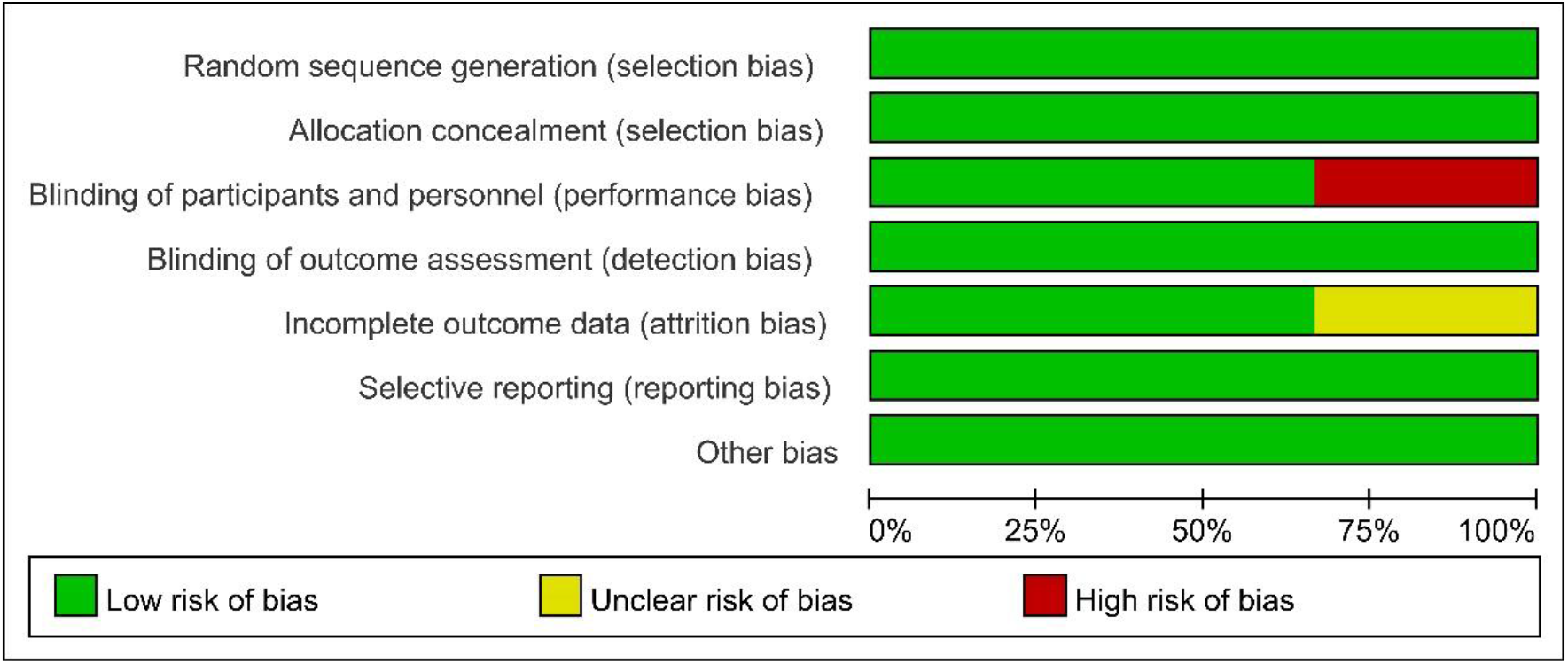
Risk of bias summary.

### Effects of interventions

#### Incidence of PEP

Three studies [17, 34, 35] have reported this result involving 1110 patients. Since significant heterogeneity was found with I^2^ >50%, we used the random-effects model. The results showed that there was no remarkable difference between patients receiving rectal NSAIDs alone and those receiving aggressive hydration in combination with NSAIDs in terms of incidence of PEP (OR, 0.43; 95% CI, 0.12-1.57; P=0.20). The forest plot was shown in **Figure 3**.

**Figure 3.**
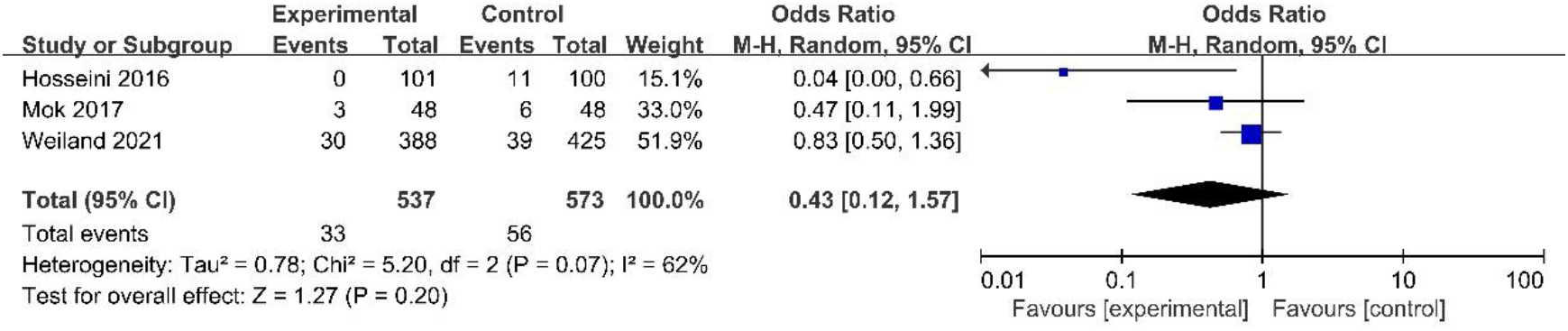
Forest plot of incidence of PEP between rectal NSAIDs alone and aggressive hydration combined with rectal NSAIDs.

#### incidence of severe PEP

Two studies [34, 35] have reported this result including 909 patients. Although I^2^ is less than 50%, the random-effects model is still applicable. The results showed that there was no remarkable difference in the incidence of severe PEP between rectal NSAIDs alone and aggressive hydration with rectal NSAIDs (OR, 0.32; 95% CI, 0.10-1.08; P=0.07). The forest plots are shown in **Figure 4**.

**Figure 4.**
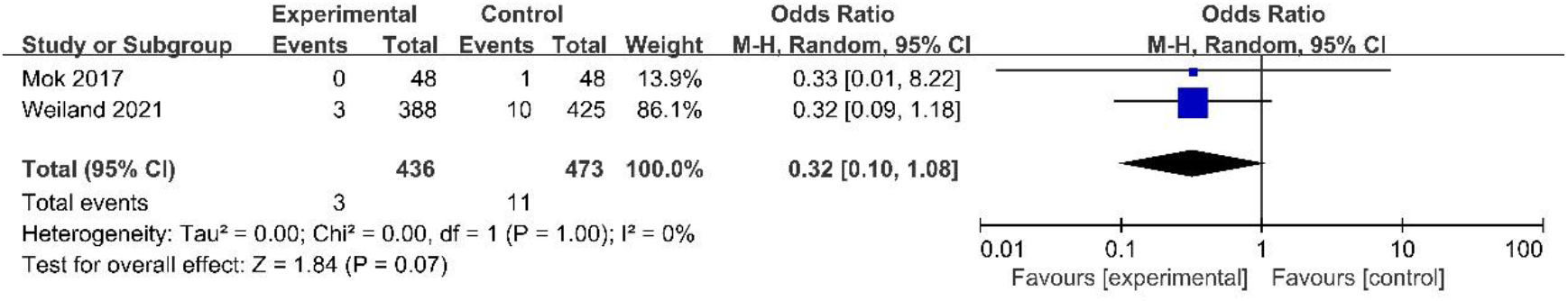
Forest plot of incidence of severe PEP between rectal NSAIDs alone and aggressive hydration combined with rectal NSAIDs.

#### Incidence of fatal PEP

Two studies [34, 35] have reported this result with 909 patients. We used the random-effects model with I2<50%. The results revealed that there was also no remarkable difference in the incidence of fatal cases between rectal NSAIDs alone and aggressive hydration combined with rectal NSAIDs (OR, 0.81; 95% CI, 0.28–2.36; P=0.70). The forest plots are shown in **Figure 5**.

**Figure 5.**
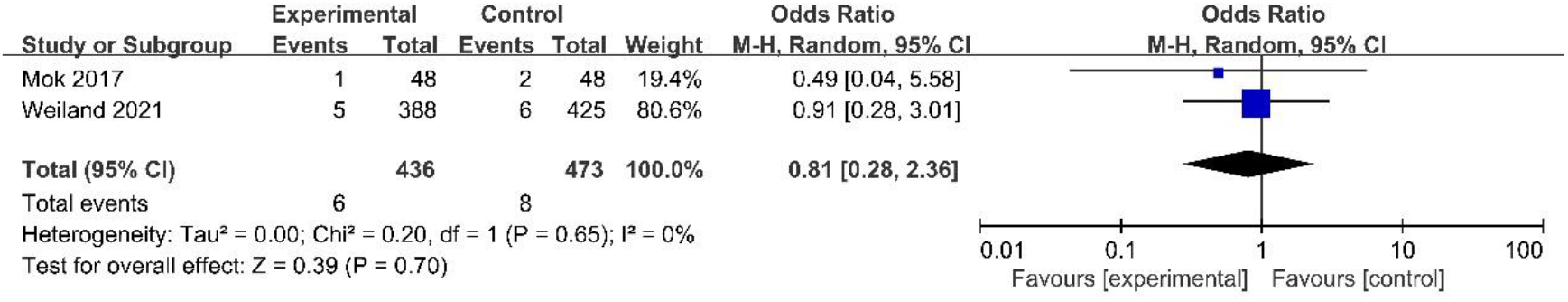
Forest plot of incidence of fatal PEP between rectal NSAIDs alone and aggressive hydration combined with rectal NSAIDs.

#### Summary of findings and quality of evidence

We evaluated the quality of evidence for the role of aggressive hydration in preventing PEP based on GRADE ratings. Evidence level for evaluating PEP prevention in the group receiving aggressive hydration versus the standard group was moderate. Evidence level for evaluating severe PEP prevention in the group receiving aggressive hydration versus the standard group was very low. Evidence level for evaluating preventive effect on fatal PEP in the group receiving aggressive hydration versus the standard group was very low. The relevant study results and GRADE recommendations are in **Table 3**.

**Table 3.**
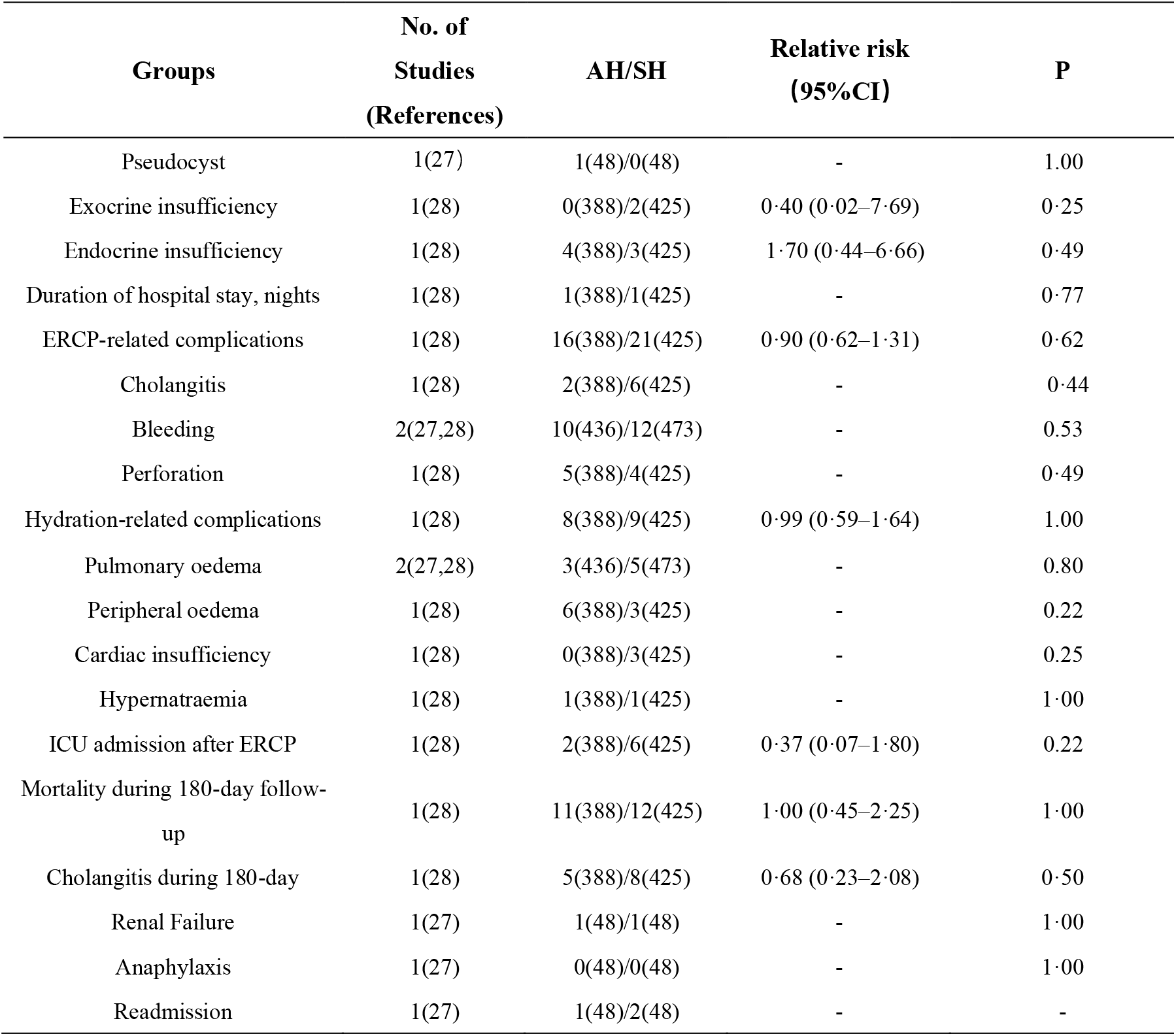
Statistical results of included studies

## Discussion

We did a meta-analysis reviewing three RCTs with 1110 patients. The results showed that the combination of prophylactic rectal NSAIDs and perioperative hydration in experimental group did not decrease the incidence of PEP compared with rectal NSAIDs alone in standard group. Meanwhile, the combination therapy had no higher effect in preventing serious cases in comparison to NSAIDs individually, with not significantly different incidence of severe PEP and fatal cases.

As an essential diagnostic and therapeutic tools of pancreaticobiliary diseases, ERCP is widely performed but also presents some complications. Among of them, exhibiting a high incidence and severity, PEP is receiving widespread attention. Although the identifying and managing the risk factors associated with ERCP can minimize the associated complications, the prevalence of PEP continues to be high. The last decade has witnessed a tremendous advance in prevention strategies for PEP, application of NSAIDs and placement of pancreatic duct stents have been broadly recognized in clinical practice for avoiding this serious complication. In particular, rectal NSAIDs, which have dropped the development of PEP significantly in 60% of cases. Moreover, a network meta-analysis including 29 studies showed that rectal NSAIDs alone presented superior results in preventing PEP compared with pancreatic duct stents alone (OR, 0.48; 95% CI, 0.26-0.87) [17]. Despite the use rectal NSAIDs, a high incidence of PEP persists due to some contraindicated populations or intolerance to NSAIDs. Recently, perioperative hydration has been recognized by several evidence-based guidance sourced form meta-analysis-level evidence, often with astonishing benefits in terms of decreased risk and improved prognosis, offering a new nonpharmacologic measure to reduce the morbidity and severity of PEP [18, 37]. A published meta-analysis including 10 RCTs with 2200 patients showed that aggressive intravenous infusion with Lactate Ringer solution was superior to standard intravenous infusion in protecting high-risk groups from PEP (OR, 0.40; 95% CI, 0.26-0.63; P < .0001) [34]. Furthermore, the positive role of aggressive hydration in decreasing the risk of developing PEP versus standard hydration was directly demonstrated in two randomized controlled trials respectively conducted by Buxbaum *et al*. (0 [0%]t of 39 patients vs 4 [17%] of 23 patients; 95% CI of difference 5.8, 35.9%; p=0.016) and Choi *et al*. (11 [4.3%] of 255 vs 25 [9.8%] of 255 patients; relative risk [RR], 0.41; 95% CI, 0.20-0.86; P = .016) [35, 38]. These studies led to international treatment guidelines recommending preoperative aggressive hydration to lower the risk of PEP [39, 40]. It has been suggested that combined aggressive hydration on the basis of rectal NSAIDs might further diminish the occurrence of PEP. However, whether there is a collaborative facilitation between rectal NSAIDs and aggressive hydration remains obscure.

Herein, we did a meta-analysis to assess the efficacy of additional aggressive hydration on patients receiving rectal NSAIDs already. Our meta-analysis showed that patients receiving aggressive fluid hydration (with Lactate Ringer solution or normal saline) plus rectal NSAIDs (100mg indomethacin or diclofenac) before or after ERCP procedure exhibited similar outcomes as patients who received rectal NSAIDs alone (or combined with restricted hydration), that is, the incidence of PEP was not significantly different (OR, 0.43; 95% CI, 0.12-1.57; P=0.20). A previous network meta-analysis presented analogous results to our study that the combination of aggressive fluid hydration with Lactate Ringer solution and rectal diclofenac 100 mg had no significant difference in preventing PEP over rectal diclofenac 100 mg alone (OR, 0·49; 95% CI, 0·26–0·94; low confidence). However, with regard to indomethacin, this meta-analysis concluded that the efficacy of the combination of standard hydration with normal saline and rectal indomethacin 100 mg surpassed rectal indomethacin 100 mg alone in preventing PEP (OR, 0·04; 95% CI, 0·00–0·66; moderate confidence) [14]. Several meta-analysis-based evidences had shown that the efficacy of indomethacin and diclofenac was similar [16, 39]. There was a high probability of false-positive results, on account of small sample sizes that included 1 RCT associated with rectal indomethacin combined standard hydration trials in this network meta-analysis. On the other hand, our results showed that no remarkable difference existed between patients receiving rectal NSAIDs alone and those receiving aggressive fluid hydration combined with rectal NSAIDs in incidence of severe PEP (OR, 0.32; 95% CI, 0.10-1.08; P=0.07) and fatal cases (OR, 0.81; 95% CI, 0.28–2.36; P=0.70). Therefore, the intravenous fluid hydration with 8-10 hours may be laborious and time-consuming for patients received rectal NSAIDs already. However, intravenous fluid infusion is still the optimal choice for patients with contraindications to NSAIDs due to its inexpensive and efficient nature.

Some shortcomings and limitations of our meta-analysis included the following: First, the number of includable literatures was not sufficient, and only three RCTs met the requirements. Secondly, sample size of some included RCTs was excessively limited, which could not exclude the possibility of false-positive or false-negative results. Third, the different intervention measures in respective RCT studies may increase the heterogeneity of our results. For instance, one RCT study [17] included in our meta-analysis compared the preventive effect of rectal indomethacin alone to the combination of intravenous normal saline and rectal indomethacin; And another RCT study [35] compared the efficacy of rectal indomethacin with intravenous Lactate Ringer solution to rectal indomethacin with intravenous normal saline in preventing PEP. However, the sensitivity analysis had shown that our final results were not affected by these different interventions.

In conclusion, aggressive perioperative hydration combined with rectal NSAID presented a modest effect in reducing the rate of PEP for patients received rectal NSAIDs already. Additionally, the combination therapy of aggressive hydration and rectal NSAIDs showed poor efficacy in preventing severe and fatal PEP. To some extent, aggressive hydration may be not necessary for patients who have received rectal NSAIDs.

## Data Availability

All data produced in the present work are contained in the manuscript.

## Statements and Declarations

There is no conflict of interest associated with this article.

